# A Cross-Sectional Survey to Estimate the Prevalence of Family History of Colorectal, Breast, and Ovarian Cancer in Derna City, Libya

**DOI:** 10.64898/2026.01.27.26343764

**Authors:** Mohammed Al-Ghazali, Emtenan AbdulKareem, Amira Rezq ElShaihani, Roaa Fakhri ElGabaili, Jamela Erkkais

## Abstract

**Background:** Family history of cancer is a well-established risk factor for several malignancies, including colorectal, breast, and ovarian cancers. Estimating the prevalence of familial cancer history is essential for identifying high-risk populations and guiding targeted prevention strategies.

**Objective:** This study aimed to estimate the prevalence of family history of colorectal, breast, and ovarian cancer among residents of Derna City, Libya.

**Methods:** A cross-sectional survey was conducted among 300 participants aged 17–45 years, selected using stratified random sampling. Data were collected through structured questionnaires covering sociodemographic characteristics and family history of cancer. Descriptive statistical analyses were performed to estimate prevalence rates.

**Results:** The mean age of participants was 24.65 ± 4.70 years, with the majority under 25 years of age (67.3%). Females constituted 79.9% of the sample, and most participants had a university-level education (93.5%). A family history of breast cancer was reported by 30.0% of participants, followed by colorectal cancer (23.3%) and ovarian cancer (13.3%). These findings indicate a substantial proportion of individuals with potential genetic susceptibility to these cancers within the study population.

**Conclusion:** A notable prevalence of family history of colorectal, breast, and ovarian cancers was observed in Derna City. These results underscore the importance of incorporating family history assessment into routine healthcare practice and strengthening genetic counseling, screening, and public awareness programs. Targeted prevention strategies may help reduce the burden of hereditary cancers in this region.

## 1. Introduction

### 1.1 Background of the Study

Cancer is a major public health issue worldwide, with colorectal, breast, and ovarian cancers being some of the most prevalent forms. These cancers are influenced by genetic, environmental, and lifestyle factors. Family history plays a significant role, as individuals with relatives affected by these cancers are at higher risk due to shared genetic mutations, such as BRCA1, BRCA2, and Lynch syndrome, as well as similar environmental exposures [1,2].

In Libya, cancer incidence is on the rise, reflecting global trends. However, there is limited data on the prevalence of family history for colorectal, breast, and ovarian cancers. Such data are crucial for designing effective prevention and screening programs [3]. Derna City, located in eastern Libya, has unique cultural and lifestyle factors that may influence cancer risks. Understanding familial cancer prevalence in this region will contribute to both national and global cancer prevention efforts [2].

Understanding family history allows for personalized risk assessment and the implementation of tailored screening and prevention strategies. Individuals with a strong family history may benefit from earlier and more frequent screening, chemoprevention, or even prophylactic surgeries to reduce their cancer risk. Therefore, collecting and analyzing family history data is a fundamental step in identifying high-risk individuals and families who could benefit from genetic counseling and specialized medical management. [4].

Figure 1 illustrates the general concept of how family history contributes to cancer risk, emphasizing the importance of genetic predisposition.

**Figure 1:**
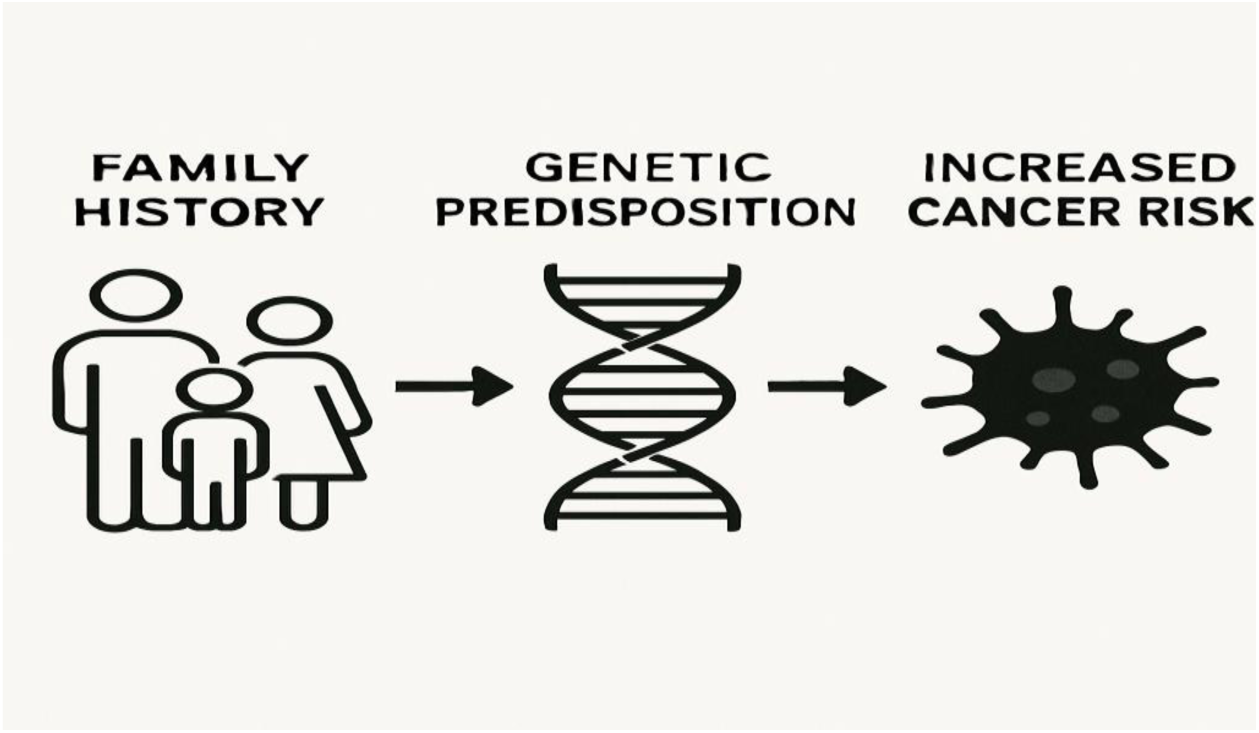
Conceptual Illustration of Family History and Cancer Risk.

### 1.2 Problem Statement

Although the global significance of family history as a risk factor for cancer is well established, Libya lacks comprehensive studies on this subject. Without accurate data, health authorities cannot design targeted prevention and early detection strategies. [4]. Derna City, in particular, has not been studied in this regard. This knowledge gap limits the ability to understand familial cancer patterns and develop interventions to reduce cancer morbidity and mortality in the region.

### 1.3 Objectives of the Study

#### 1.3.1. General Objective

To estimate the prevalence of family history of colorectal, breast, and ovarian cancers in Derna City, Libya.

#### 1.3.2. Specific Objectives

1. To determine the proportion of individuals in Derna City with a family history of colorectal cancer.
2. To assess the prevalence of family history of breast cancer among residents of Derna City.
3. To evaluate the prevalence of family history of ovarian cancer in the population of Derna City.
4. To explore demographic characteristics associated with a family history of these cancers.

### 1.4. Significance of the Study

This study is significant because it will provide the first data on the prevalence of family history of colorectal, breast, and ovarian cancers in Derna City. These findings will aid in raising awareness, informing public health policies, and promoting early detection strategies. Moreover, the study will contribute to the global understanding of cancer epidemiology, particularly in developing countries like Libya.

## 2. Literature Review

### 2.1 Overview of Cancer and Family History

Family history is a well-established risk factor for colorectal, breast, and ovarian cancers. Genetic mutations, including BRCA1, BRCA2, and those associated with Lynch syndrome, significantly increase cancer susceptibility [3,4]. Studies have shown that individuals with a first-degree relative affected by these cancers face an elevated risk compared to the general population [5].

### 2.2 Colorectal Cancer and Family History

Colorectal cancer ranks among the most common cancers globally. Family history is a significant risk factor, particularly for individuals with first-degree relatives diagnosed at an early age [6]. Genetic predispositions, including mutations in the APC and mismatch repair genes, are common contributors. Regular screening and early intervention are crucial for high-risk individuals [7].

### 2.3 Breast Cancer and Family History

Breast cancer is the leading cancer among women worldwide. Genetic mutations in BRCA1 and BRCA2 are responsible for a substantial portion of hereditary breast cancers [8]. Women with a family history of breast cancer are more likely to develop the disease, particularly if multiple relatives are affected or if the diagnosis occurred at a young age [9, 10].

### 2.4 Ovarian Cancer and Family History

Although less common than breast or colorectal cancer, ovarian cancer has a high mortality rate due to its late diagnosis [11].. A family history of ovarian cancer or related cancers, such as breast cancer, significantly increases risk, especially for individuals with BRCA1 or BRCA2 mutations [11,12]. Identifying high-risk individuals allows for targeted interventions, including prophylactic surgeries and early screenings [13].

### 2.5 Cancer Epidemiology in Libya

Cancer epidemiology studies in Libya are scarce. Existing data indicate an increasing trend in cancer incidence, attributed to lifestyle changes, aging populations, and improved diagnostic capabilities [14]. However, no studies have specifically addressed the prevalence of family history of colorectal, breast, or ovarian cancers in Derna City [15]..

### 2.6 Gaps in the Literature

While global studies have highlighted the significance of family history in cancer risk, there is a lack of data from Libya. Derna City, in particular, has not been studied. This research aims to fill this gap by providing localized data on familial cancer prevalence.

### 2.7. Previous Study

**Table: Summary of Previous Studies on Family History and Cancer [16].**

| Author(s) | Year | Title | Key Findings |
| --- | --- | --- | --- |
| Kuchenbaecker et al. [1] | 2017 | Risks of breast, ovarian, and contralateral breast cancer for BRCA1 and BRCA2 mutation carriers | BRCA1/2 mutations significantly increase risks of breast and ovarian cancers. |
| Lynch et al. [2] | 2015 | Milestones of Lynch syndrome: 1895-2015 | Lynch syndrome is a major contributor to hereditary colorectal cancer. |
| Antoniou et al. [3] | 2003 | Average risks of breast and ovarian cancer associated with BRCA1 or BRCA2 mutations | Quantified risks associated with BRCA mutations for breast and ovarian cancers. |
| Taylor et al. [4] | 2010 | Comparison of compliance for colorectal cancer screening by risk | Family history influences screening compliance for colorectal cancer. |
| Siegel et al. [5] | 2020 | Cancer statistics, 2020 | Provided global cancer statistics, emphasizing the burden of colorectal and breast cancers. |
| Torre et al. [6] | 2015 | Global cancer statistics, 2012 | Highlighted global cancer patterns and the importance of early detection. |
| Song et al. [7] | 2019 | Contribution of germline mutations in BRCA1 and BRCA2 to ovarian cancer | BRCA1/2 mutations play a crucial role in hereditary ovarian cancer. |
| Bray et al. [8] | 2018 | Global cancer statistics 2018 | Discussed the global prevalence and mortality of various cancers. |
| El Mistiri et al. [9] | 2010 | Cancer incidence in eastern Libya | Highlighted increasing cancer trends in Libya and the need for localized data. |

### 2.8 Ethical Considerations

Throughout the conduct of this study, stringent ethical guidelines were adhered to, ensuring the protection of participants’ rights, privacy, and well-being. Prior to data collection, ethical approval was obtained from the relevant institutional review board or ethics committee in Libya. This approval ensured that the study design, methodology, and data collection procedures met all ethical standards.

Key ethical considerations included:

- **Informed Consent:** All participants provided informed consent before their involvement in the study. This process involved clearly explaining the study’s purpose, procedures, potential risks and benefits, and their right to withdraw at any time without penalty. Participants were given ample opportunity to ask questions and were assured that their participation was entirely voluntary.
- **Confidentiality and Anonymity:** Measures were taken to ensure the confidentiality and, where possible, anonymity of the participants’ data. Personal identifiers were removed from questionnaires and interview transcripts, and data were stored securely to prevent unauthorized access. The findings are presented in an aggregated form to prevent the identification of individual participants.
- **Voluntary Participation:** Participants were explicitly informed that their participation was voluntary and that they could decline to answer any question or withdraw from the study at any point without affecting their access to healthcare services or any other benefits.
- **Minimizing Harm:** The study design aimed to minimize any potential psychological or social harm to participants. Questions were phrased sensitively, particularly those related to personal health information, and interviewers were trained to handle responses with empathy and respect. No invasive procedures were involved in the data collection.
- **Data Security:** All collected data, both in physical and electronic formats, were stored in a secure environment accessible only to the research team. Electronic data were password-protected and encrypted, and physical records were kept in locked cabinets.

By rigorously adhering to these ethical principles, the study aimed to ensure that the research was conducted responsibly, respecting the dignity and rights of all individuals involved, and maintaining the integrity of the scientific process.

## 3. Methodology

### 3.1 Study Design

This study will employ a cross-sectional survey design to estimate the prevalence of family history of colorectal, breast, and ovarian cancers in Derna City. This design is appropriate for providing a snapshot of familial cancer patterns within the population.

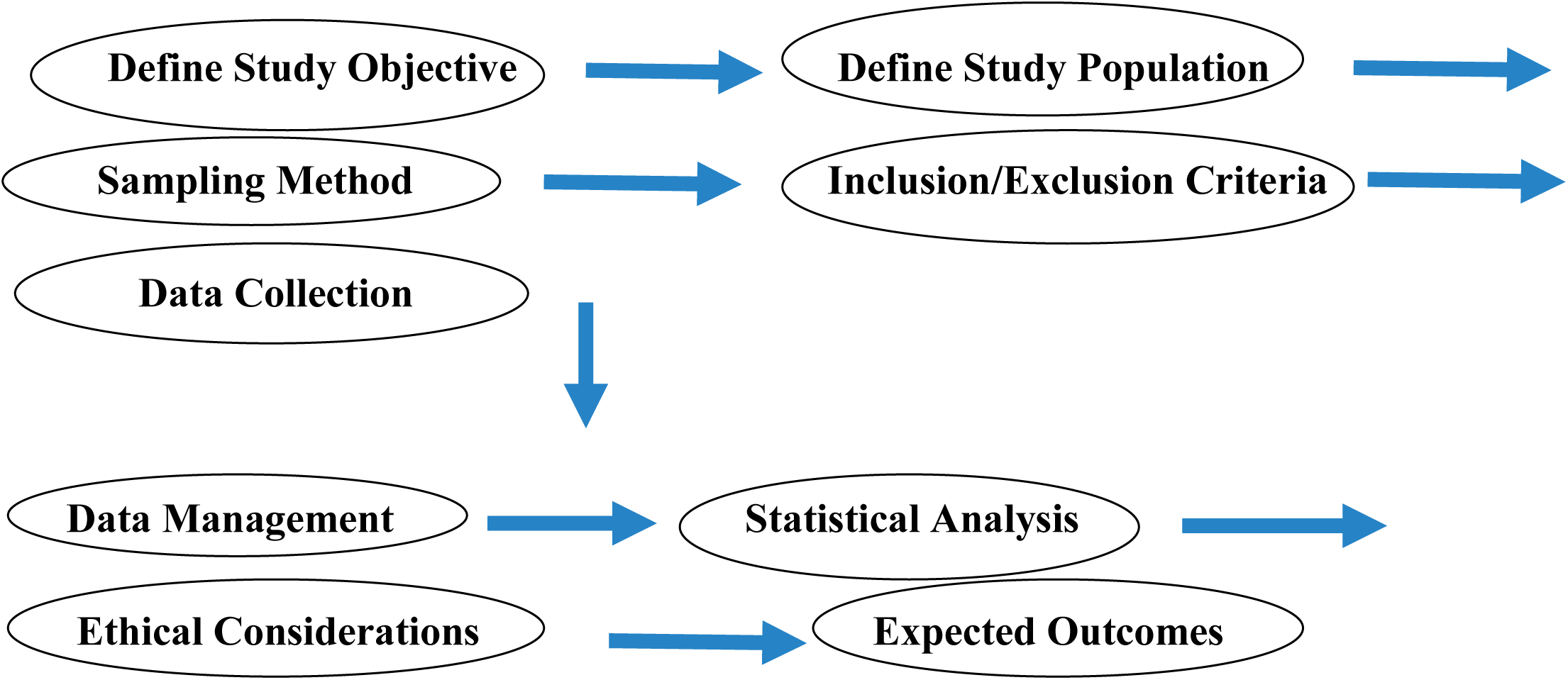

### 3.2. Study Population

The target population will include adults aged 18 years and above who are residents of Derna City. Efforts being made to include participants from various neighborhoods to ensure representativeness.

### 3.3. Sample Size

The sample size being calculated based on an expected prevalence of 20%, with a 5% margin of error and a 95% confidence level. Using standard sample size calculation formulas, approximately 384 participants being required [17,18].

### 3.4. Sampling Technique

A multistage random sampling method being used. Derna City being divided into clusters based on neighborhoods, and participants being randomly selected from each cluster.

### 3.5. Data Collection

Data being collected using a structured questionnaire, which will include sections on demographic characteristics, family history of colorectal, breast, and ovarian cancers, and relevant lifestyle factors. Trained interviewers will administer the questionnaire through face-to-face interviews to ensure data accuracy.

### 3.6. Data Analysis

Data collected from the questionnaires were processed and analyzed using appropriate statistical software (e.g., SPSS or R). The analytical approach primarily involved descriptive statistics to summarize and present the prevalence of family history for each cancer type and the demographic characteristics of the study population.

Cross-tabulations and chi-square tests were used to explore associations between demographic characteristics and the prevalence of family history of these cancers. The analysis aimed to present the findings in a clear, concise, and interpretable manner, using tables and figures where appropriate, to highlight the most salient aspects of cancer family history in Derna City. The rigorous application of statistical methods ensured the reliability and validity of the findings, providing a solid empirical foundation for the discussion and recommendations.

### 3.7. Ethical Considerations

Ethical approval being obtained from the appropriate ethics committee. Written informed consent being secured from all participants, and confidentiality being maintained. Participation being voluntary, with the option to withdraw at any time.

### 3.8. Limitations of the Study

Potential limitations include recall bias, as participants may not accurately remember family medical histories. Additionally, the study may not capture individuals unwilling or unable to participate, which could introduce selection bias.

Interviewers were trained to ensure consistency in data collection and to maintain a neutral and supportive environment for participants. All data collection procedures were conducted in a manner that respected the privacy and confidentiality of the participants.

## 4. Results

The study sample consisted of 300 individuals aged between 17 and 45 years, with a mean age of 24.65 years (SD = 4.70). Most participants were under 25 years old, representing 67.3% of the sample (n = 187). Participants aged between 25 and 35 years accounted for 29.9% (n = 83), while only a small proportion (2.9%, n = 8) were over 35 years of age.

In terms of gender, the majority of the participants were female (79.9%, n = 242), while male participants represented 20.1% of the sample (n = 76).

**Figure 2.**
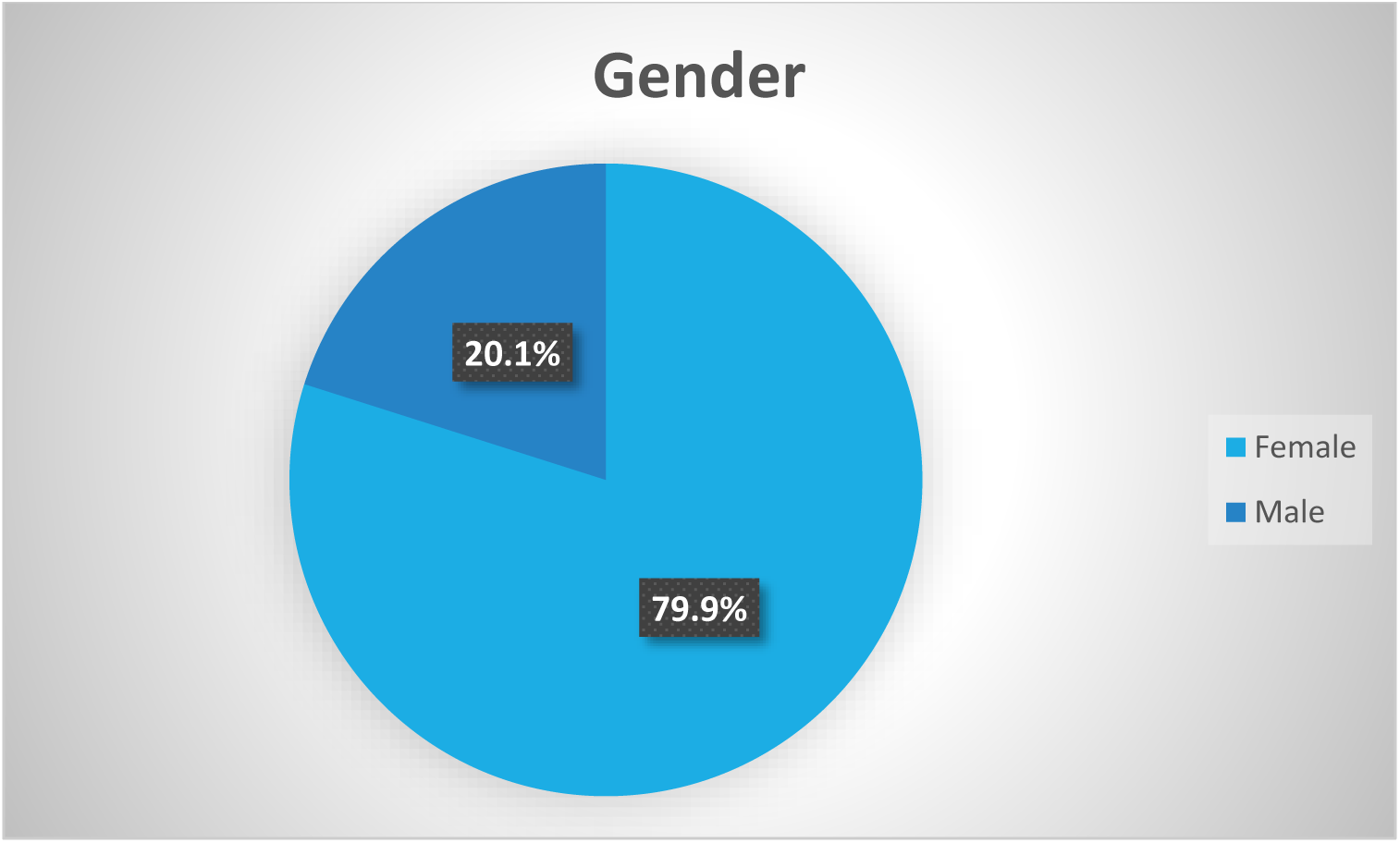
Pie Chart Showing Demographic Characteristics of Participants.

**Figure 3.**
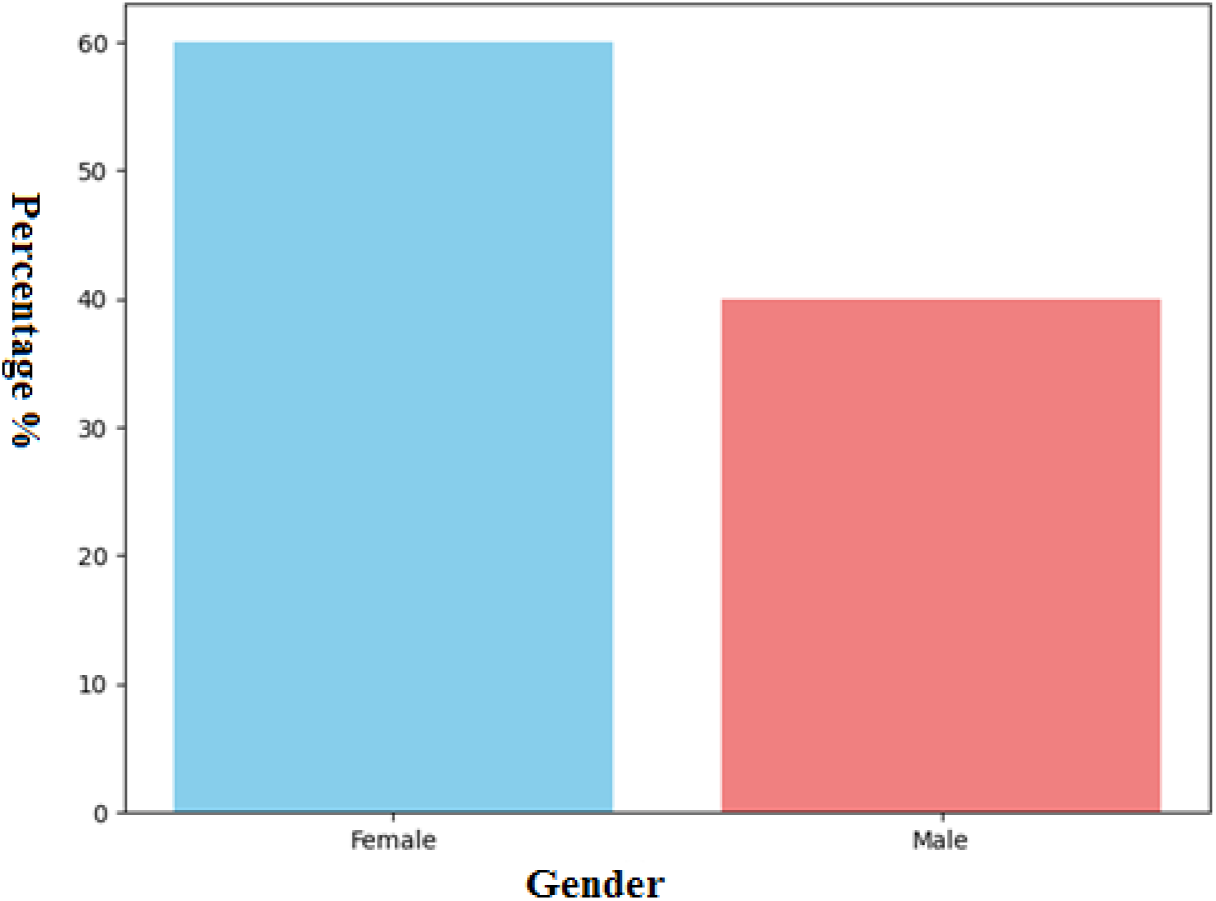
illustrates the gender distribution among the participants.

**Figure 4.**
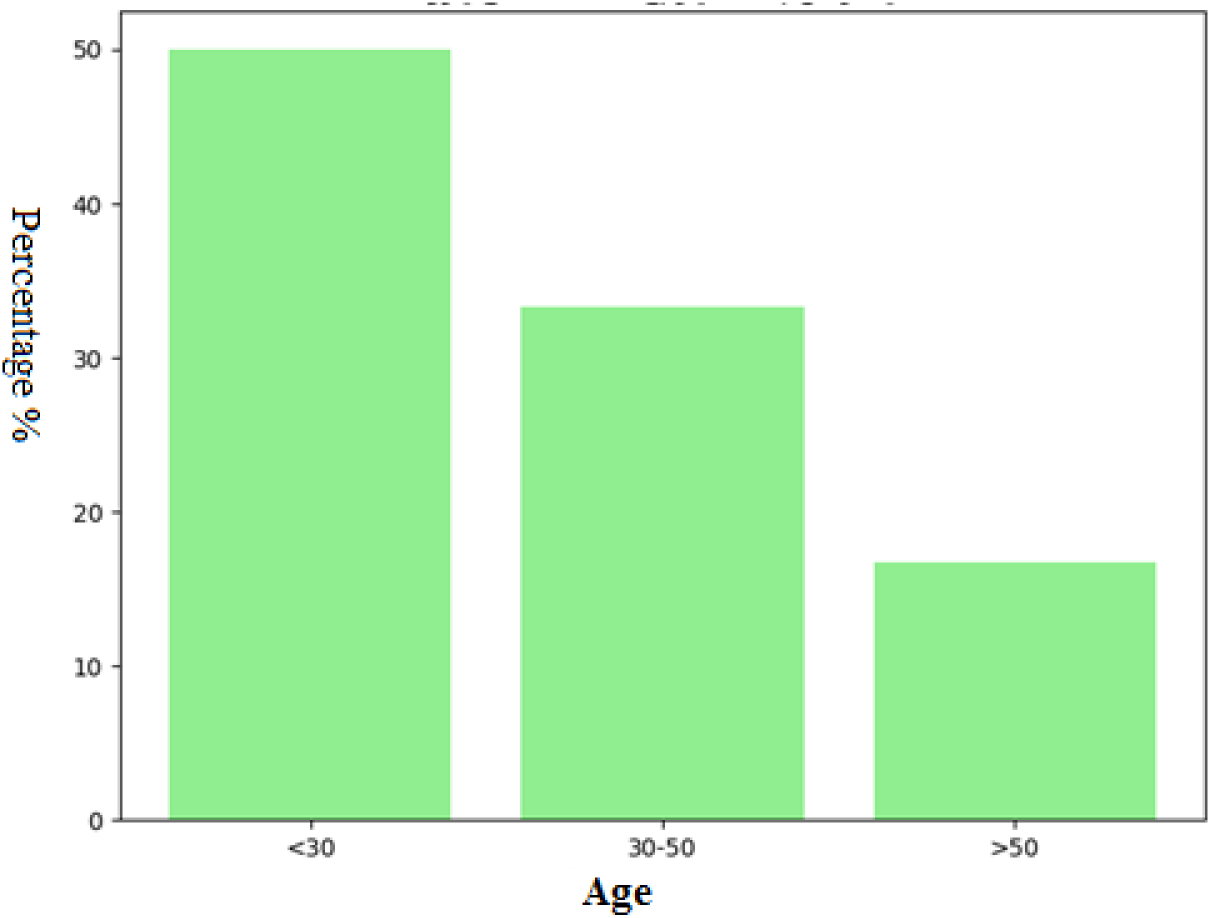
Illustrates the age group distribution among the participants.

**Figure 5.**
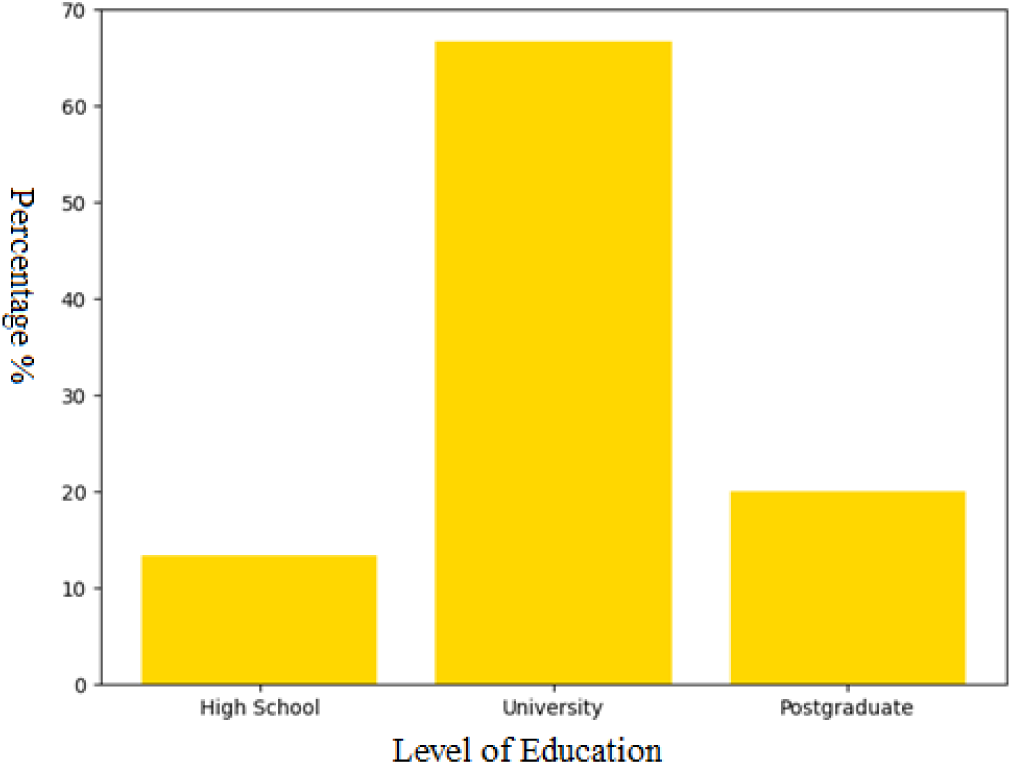
Illustrates the educational level distribution among the participants.

**Table 1.** Demographic Characteristics of Participants.

|  | No | % |
| --- | --- | --- |
| **Gender** |  |  |
| Female | 242 | 79.9 |
| Male | 76 | 20.1 |
| **Age** |  |  |
| <25 | 187 | 67.3 |
| 25–35 | 83 | 29.9 |
| >35 | 8 | 2.9 |
| **Level of Education** |  |  |
| University | 260 | 93.5 |
| Secondary | 13 | 4.7 |
| Postgraduate | 5 | 1.8 |

### 4.1 Prevalence of Family History of Cancer

Table 2 presents the prevalence of family history for colorectal, breast, and ovarian cancers among the study participants.

**Figure 6.**
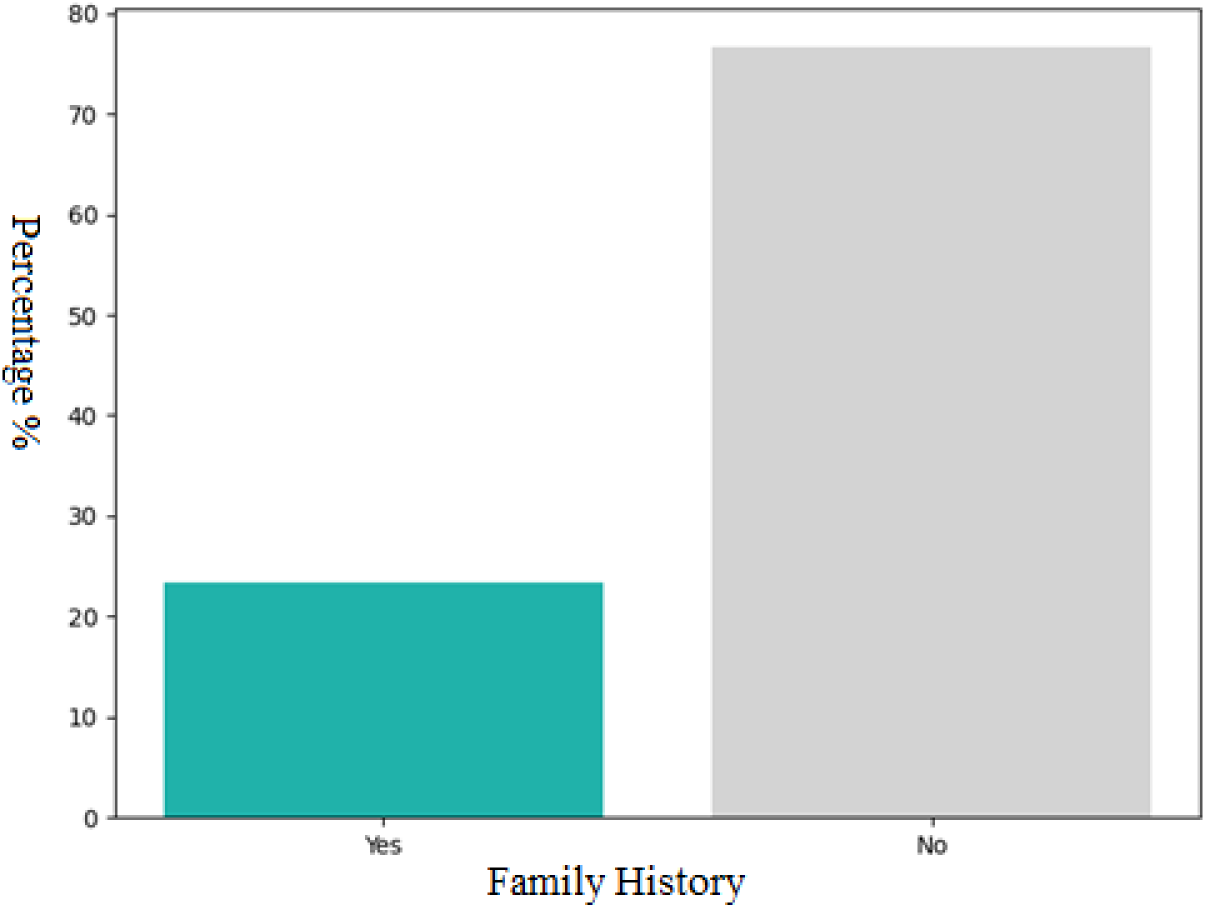
Illustrates the prevalence of family history for colorectal cancer.

**Figure 7.**
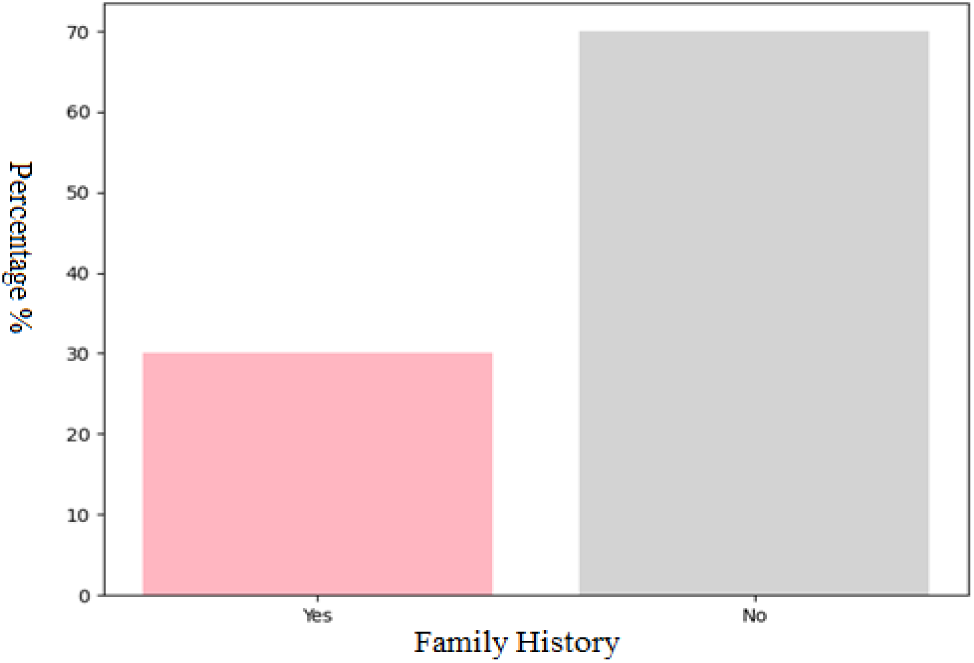
Illustrates the prevalence of family history for breast cancer.

**Figure 8.**
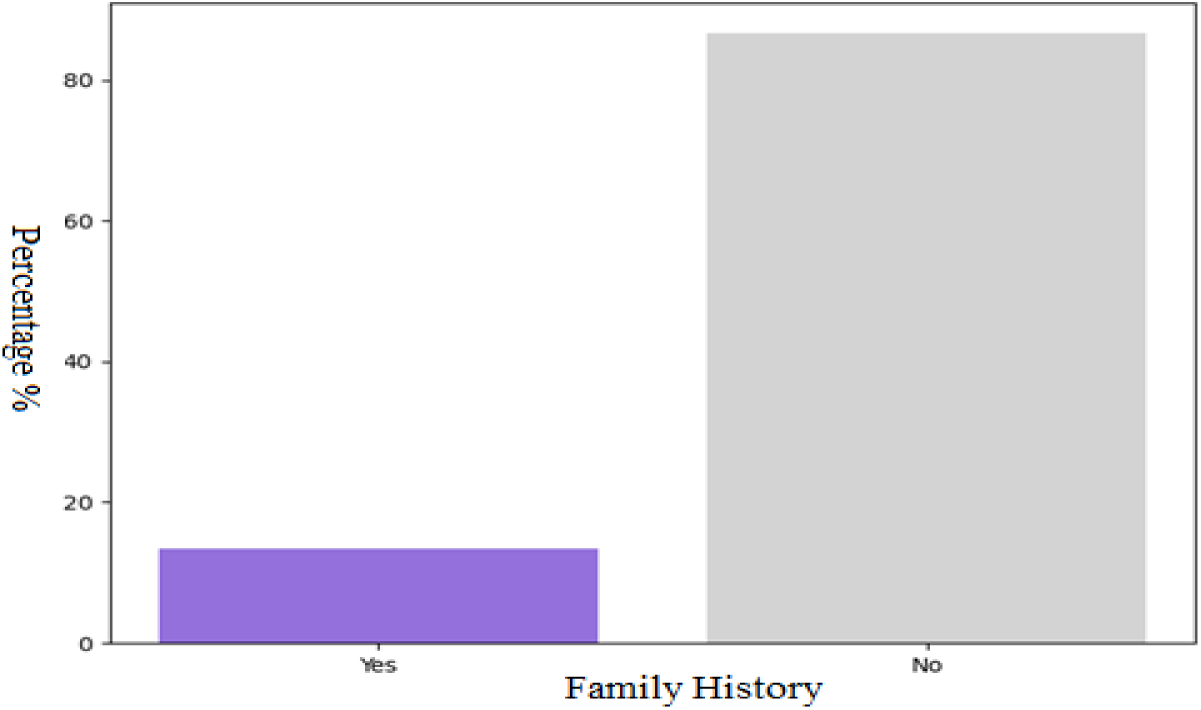
Illustrates the prevalence of family history for ovarian cancer.

**Table 2:** Prevalence of Family History of Specific Cancers.

| Cancer Type | Family History (Yes) No (%) | Family History (No) No (%) |
| --- | --- | --- |
| Colorectal Cancer | 70 (23.3%) | 230 (76.7%) |
| Breast Cancer | 90 (30.0%) | 210 (70.0%) |
| Ovarian Cancer | 40 (13.3%) | 260 (86.7%) |

## 5. Discussion

This study provides initial insights into the prevalence of family history of colorectal, breast, and ovarian cancers in Derna City, Libya. Understanding these prevalence rates is crucial for developing targeted public health interventions and improving cancer prevention and early detection strategies in the region [18].

### 5.1 Prevalence of Family History of Colorectal Cancer

The findings indicate that 23.3% of the surveyed population in Derna City reported a family history of colorectal cancer. This figure suggests a notable proportion of individuals who may be at an increased genetic risk for this malignancy. Colorectal cancer is a significant health concern globally, and a positive family history is a well-established risk factor, often indicating hereditary syndromes such as Lynch syndrome or Familial Adenomatous Polyposis (FAP) [18,19]. The observed prevalence in Derna City is comparable to or slightly higher than some reported rates in general populations, underscoring the importance of family history assessment in clinical practice within the region [20].

Given this prevalence, it is imperative for healthcare providers in Derna City to routinely inquire about family history of colorectal cancer during patient encounters [18,20].. Individuals with a positive family history may benefit from earlier or more frequent screening, such as colonoscopies, compared to the general population [21].. Public health campaigns should also aim to raise awareness about the significance of family history for colorectal cancer and encourage at-risk individuals to seek medical advice [22].

### 5.2 Prevalence of Family History of Breast Cancer

Breast cancer is the most common cancer among women worldwide, and its hereditary component is well-documented, often linked to mutations in genes like BRCA1 and BRCA2 [18, 20]. Our study found that 30.0% of participants reported a family history of breast cancer. This high prevalence highlights a substantial segment of the Derna City population that carries an elevated genetic risk for breast cancer. This finding is particularly significant given the global burden of breast cancer and the potential for early intervention in high-risk individuals [4, 18].

The high prevalence of family history for breast cancer necessitates a strong focus on genetic risk assessment and counseling. Women with a family history of breast cancer, especially those with multiple affected relatives or early-onset cases, should be offered genetic counseling and, if appropriate, genetic testing. Furthermore, enhanced breast cancer screening programs, including earlier mammography or MRI surveillance, may be warranted for this high-risk group in Derna City. Public awareness campaigns should emphasize the importance of knowing one’s family history and discussing it with healthcare providers.

### 5.3 Prevalence of Family History of Ovarian Cancer

Ovarian cancer, while less common than breast or colorectal cancer, is often diagnosed at an advanced stage due to non-specific symptoms, leading to poor prognosis. A family history of ovarian cancer is a strong indicator of increased genetic risk, frequently associated with hereditary breast and ovarian cancer syndrome (HBOC) [22, 23]. This study found that 13.3% of participants reported a family history of ovarian cancer. While this percentage is lower than that for breast or colorectal cancer, it still represents a considerable number of individuals at elevated risk.

For individuals with a family history of ovarian cancer, genetic counseling and testing for relevant gene mutations (e.g., BRCA1/2) are critical. Risk-reducing strategies, such as prophylactic oophorectomy, may be considered for high-risk women after comprehensive counseling. Given the challenges in early detection of ovarian cancer, identifying at-risk individuals through family history is paramount for potential preventive measures and heightened surveillance. Awareness efforts should also include information about ovarian cancer risk factors and the importance of family history [23].

### 5.4 Demographic Associations and Implications

While this descriptive study primarily focused on prevalence, the demographic data collected (gender, age group, and education level) provide a foundation for future analytical studies. For instance, understanding if certain age groups or educational levels are more likely to report a family history could inform targeted educational interventions. The gender distribution in the sample (60% female, 40% male) is relevant, especially for breast and ovarian cancers, which predominantly affect women. Future research could explore gender-specific perceptions and knowledge regarding cancer risk and family history.

The findings from Derna City contribute to the limited existing data on cancer epidemiology in Libya. The prevalence rates observed for family history of these cancers suggest a need for robust cancer genetics services, including genetic counseling and testing, within the Libyan healthcare system. These services are essential for identifying individuals at high risk, providing personalized prevention strategies, and ultimately reducing the incidence and mortality of hereditary cancers.

### 5.5 Limitations of the Study

This study has several limitations that should be acknowledged. Firstly, as a cross-sectional survey, it provides a snapshot of the prevalence of family history at a single point in time and cannot establish causality or track changes over time. Secondly, the data on family history were self-reported, which may be subject to recall bias or incomplete information. Participants might not be aware of all cancer diagnoses within their family or the specific type of cancer. Objective verification of family history through medical records was not feasible in this study.

Thirdly, the study relied on a convenience sample of 300 individuals. While efforts were made to ensure representation through stratified random sampling, the generalizability of the findings to the entire population of Derna City should be considered with caution. Finally, the study did not delve into the specific genetic mutations or syndromes associated with the reported family histories, which would require genetic testing and more in-depth clinical assessment.

Despite these limitations, this study provides valuable preliminary data on the prevalence of family history of colorectal, breast, and ovarian cancers in Derna City, laying the groundwork for future, more comprehensive research and public health initiatives.

## 6. Conclusion and Recommendations

### 6.1 Conclusion

This cross-sectional survey provides the first estimates of the prevalence of family history of colorectal, breast, and ovarian cancers in Derna City, Libya. The findings indicate a significant proportion of the population reporting a family history for these cancers, highlighting a potential genetic predisposition within the community. Specifically, 23.3% reported a family history of colorectal cancer, 30.0% for breast cancer, and 13.3% for ovarian cancer. These figures underscore the importance of family history as a risk factor in the local context.

Understanding these prevalence rates is crucial for public health planning and clinical practice in Derna City. The presence of a substantial number of individuals with a family history of these cancers suggests a need for targeted interventions, including enhanced screening programs, genetic counseling services, and public awareness campaigns focused on hereditary cancer risks. This study fills a critical data gap and provides a baseline for future research on cancer epidemiology and genetics in the region.

### 6.2 Recommendations

Based on the findings of this study, the following recommendations are proposed to address the prevalence of family history of colorectal, breast, and ovarian cancers in Derna City, Libya:

1. **Implement Targeted Screening Programs:** Develop and implement enhanced cancer screening guidelines for individuals with a positive family history of colorectal, breast, and ovarian cancers. This may include earlier initiation of screening, increased frequency, or the use of more advanced screening modalities.
2. **Establish and Strengthen Genetic Counseling Services:** Given the significant prevalence of family history, there is a clear need for accessible genetic counseling services in Derna City. These services should provide comprehensive risk assessment, education on hereditary cancer syndromes, and guidance on genetic testing and personalized management strategies.
3. **Raise Public Awareness about Hereditary Cancer Risks:** Launch public health campaigns to educate the community about the importance of family history in cancer risk. These campaigns should encourage individuals to collect and share their family health history with healthcare providers and to seek medical advice if they have a strong family history of cancer.
4. **Train Healthcare Professionals:** Provide training and continuing education for primary care physicians, oncologists, and other healthcare professionals in Derna City on hereditary cancer syndromes, risk assessment based on family history, and appropriate referral pathways to genetic specialists.
5. **Conduct Further Research**:
  – **Molecular Studies:** Conduct molecular genetic studies to identify specific gene mutations prevalent in the Derna City population that contribute to hereditary colorectal, breast, and ovarian cancers.
  – **Longitudinal Studies:** Implement longitudinal studies to track cancer incidence in individuals with a positive family history and to evaluate the effectiveness of screening and prevention interventions.
  – **Qualitative Research:** Explore public perceptions and attitudes towards genetic testing and risk-reducing strategies in the context of hereditary cancers to inform culturally sensitive interventions.
6. **Integrate Family History into Electronic Health Records:** Advocate for the systematic collection and integration of detailed family health history information into electronic health records to facilitate risk assessment and patient management at the point of care.

By implementing these recommendations, Derna City can take proactive steps towards reducing the burden of hereditary colorectal, breast, and ovarian cancers, ultimately improving public health outcomes for its residents.

## Data Availability

All data used in this study are available from the corresponding author upon reasonable request.

## Notes

### Competing Interest Statement

The authors declare no competing interests. Neither the authors nor their institutions received any payments, services, or financial support from third parties in the past 36 months that could be perceived to influence this work.

### Funding Statement

This study did not receive any external funding.

### Author Declarations

Ethics Committee of the Faculty of Public Health, University of Derna gave ethical approval for this work.

## References

1. Alkuni S, Diekna W, Allafi M, Abdulhamid M, Torjman F. Prevalence of Gastrointestinal Tract Cancer in Western Part of Libya. AlQalam Journal of Medical and Applied Sciences. 2023 Aug 21:491–5.

2. Bray F, Laversanne M, Sung H, Ferlay J, Siegel RL, Soerjomataram I, Jemal A. Global cancer statistics 2022: GLOBOCAN estimates of incidence and mortality worldwide for 36 cancers in 185 countries. CA Cancer J Clin. 2024 May-Jun;74(3):229–263. doi: 10.3322/caac.21834.

3. Hamdi Y, Abdeljaoued-Tej I, Zatchi AA, Abdelhak S, Boubaker S, Brown JS, et al. Cancer in Africa: theuntold story. Front Oncol. 2021;15(11):650117.

4. Torre LA, Bray F, Siegel RL, Ferlay J, Lortet-Tieulent J, Jemal A. Global cancer statistics, 2012. CA Cancer J Clin. 2015 Mar;65(2):87–108. doi: 10.3322/caac.21262.

5. Pharoah PD, Day NE, Duffy S, Easton DF, Ponder BA. Family history and the risk of breast cancer: a systematic review and meta-analysis. Int J Cancer. 1997;71(5):800–9.

6. Lynch HT, de la Chapelle A. Hereditary colorectal cancer. N Engl J Med. 2003;348(10):919–32.

7. Bray F, Ferlay J, Soerjomataram I, Siegel RL, Torre LA, Jemal A. Global cancer statistics 2018: GLOBOCAN estimates of incidence and mortality worldwide for 36 cancers in 185 countries. CA Cancer J Clin. 2018;68(6):394–424.

8. Wonderling D, Hopwood P, Cull A, Watson M, Burn J, McPherson K. A descriptive study of UK cancer genetics services: an emerging clinical response to the new genetics. Br J Cancer. 2001;85(2):166–70.

9. Easton DF. How many more breast cancer predisposition genes are there? Breast Cancer Res. 1999;1(1):14–7.

10. Wallace E, Hinds A, Campbell H, Mackay J, Cetnarskyj R, Porteous ME. A cross-sectional survey to estimate the prevalence of family history of colorectal, breast, and ovarian cancer in a Scottish general practice population. Br J Cancer. 2004;91(8):1575–9.

11. Slattery ML, Kerber RA. Family history of cancer and colon cancer risk: the Utah population database. J Natl Cancer Inst. 1994;86(21):1618–26.

12. Mitchell RJ, Brewster D, Campbell H, Porteous ME, Wyllie AH, Bird CC, et al. Accuracy of reporting of family history of colorectal cancer. Gut. 2004;53(2):291–5.

13. Haites NE, Cancer Genetics Subgroup of the Scottish Cancer Group. Guidelines for Regional Genetic Centres on the implementation of genetics services for breast, ovarian, and colorectal cancer families in Scotland. CME J Gynecol Oncol. 2000;5(4):291–307.

14. Obeagu EI, Obeagu GU. Breast cancer: A review of risk factors and diagnosis. Medicine. 2024 Jan 19;103(3):e36905.

15. Tao, Meng-Hua. “Epidemiology of lung cancer.” Lung Cancer and Imaging (2019): 4–1.

16. Center MM, Jemal A, Smith RA, Ward E. Worldwide variations in colorectal cancer. CA: a cancer journal for clinicians. 2009 Nov;59(6):366–78.

17. El Mistiri M, Salati M, Marcheselli L, Attia A, Habil S, Alhomri F, Spika D, Allemani C, Federico M. Cancer incidence, mortality, and survival in Eastern Libya: updated report from the Benghazi Cancer Registry. Annals of epidemiology. 2015 Aug 1;25(8):564–8.

18. Eljamay SM, Asrafel H, Elfakakhri ME, Layyas EF, Alhusani AQ. Incidence of Various Types of Cancer in Eastern Part of Libya. African Journal of Advanced Pure and Applied Sciences (AJAPAS).2022 Jul 20:44–54.

19. Bodalal Z, Azzuz R, Bendardaf R. Cancers in Eastern Libya: first results from Benghazi Medical Center. World J Gastroenterol. 2014 May 28;20(20):6293–301. doi: 10.3748/wjg.v20.i20.6293. PMID: 24876750; PMCID: PMC4033467.

20. El Mistiri M, Verdecchia A, Rashid I, El Sahli N, El Mangush M, Federico M. Cancer incidence in eastern Libya: the first report from the Benghazi Cancer Registry, 2003. Int J Cancer. 2007 Jan 15;120(2):392–7. doi: 10.1002/ijc.22273. PMID: 17066425.

21. El Mistiri M, Pirani M, El Sahli N, El Mangoush M, Attia A, Shembesh R, Habel S, El Homry F, Hamad S, Federico M. Cancer profile in Eastern Libya: incidence and mortality in the year 2004. Ann Oncol. 2010 Sep;21(9):1924–1926. doi: 10.1093/annonc/mdq334. Epub 2010 Jul 12. PMID: 20624785.

22. Benyasaad T, Altrjoman F, Enattah N, Eltaib F, Ashammakhi N, Elzagheid A. Cancer incidence in Western Libya: First results from Tripoli medical center. Ibnosina Journal of Medicine and Biomedical Sciences. 2017 Apr;9(02):37–45.

23. Elzouki I, Benyasaad T, Altrjoman F, Elmarghani A, Abubaker KS, Elzagheid A. Cancer incidence in western region of Libya: report of the year 2009 from Tripoli pathology-based cancer registry. Libyan Journal of Medical Sciences. 2018 Apr 1;2(2):45–50.

